# Understanding the Meaning of a Good Death for People living with Parkinson’s Disease: Qualitative study

**DOI:** 10.1101/2025.11.17.25340426

**Authors:** Leonardo Martins, Rasa Mikelyte, Ricardo Silva Carvalho, Henrique Ballalai Ferraz, Déborah Oliveira, Julia Maria Vanelli, Natália Rocha Tardelli, Fernanda Bono Fukushima, Edison Iglesias de Oliveira Vidal

**Affiliations:** Public Health Department, Medical School Botucatu, São Paulo State University (UNESP), Botucatu, Brazil; Centre for Health Services Studies, University of Kent, Canterbury, United Kingdom; Internal Medicine Department, Medical School Botucatu, São Paulo State University (UNESP), Botucatu, Brazil; Movement Disorders Division, Neurology Department, Federal University of São Paulo, São Paulo, Brazil; Universidad Andrés Bello, Faculty of Nursing, Campus Viña del Mar, Chile, and Millenium Institute for Care Research (MICARE); Geriatrics Division, Internal Medicine Department, Medical School Botucatu, São Paulo State University (UNESP), Botucatu, Brazil; Pain and Palliative Care Division, Surgical Specialties Department, Medical School Botucatu, São Paulo State University (UNESP), Botucatu, Brazil

**Keywords:** Parkinson’s disease, Palliative care, Death, Qualitative Studies

## Abstract

**Background and Objectives:** Parkinson’s disease is the second most common neurodegenerative disorder globally. Despite its prevalence, the provision of Palliative Care for people living with Parkinson’s disease (PLwPD) is often delayed or entirely absent. To date, no study has explored what constitutes a "good death", a central goal of palliative care, from the perspective of PLwPD themselves. We aimed to give voice to PLwPD on this topic through a qualitative approach.

**Methods:** In this cross-sectional multicenter qualitative study, we conducted semi-structured interviews with 30 PLwPD selected through purposive sampling from four geriatric and neurology outpatient clinics between May 2021 and December 2022. An interdisciplinary team analyzed the transcripts using inductive thematic analysis. The process involved independent coding by three researchers, followed by iterative collaborative team discussions to refine and standardize the analysis, all grounded in a constructionist paradigm. To ensure methodological rigor, we employed techniques of triangulation, thick descriptions, and reflexivity.

**Results:** The sample was diverse in terms of race/ethnicity, gender, age (36–84 years), religious affiliation, educational background, and disease stage. We identified two major themes: *Fears* and *Coping*. Reported fears included experiencing disability, pain and discomfort, fear of feeling shame, fear of being a burden, fear of being abandoned and left helpless. Coping was a multidimensional theme, comprising the relational experience of feeling well cared for (defined by being valued, receiving clear and honest communication, and being treated with love and kindness) alongside the active strategies of finding opportunities for joy and drawing on religiosity and spirituality. Religiosity/spirituality appeared as a key factor in emotional regulation, fostering a sense of purpose and acceptance in the face of death.

**Discussion:** Our findings suggest that improving palliative care for PLwPD requires an approach that actively addresses specific fears and strengthens the multiple dimensions of coping, which includes fostering opportunities for joy, supporting spirituality, and enhancing the relational experience of feeling well cared for. This study illuminates often-overlooked aspects of care and provides a basis for the development of person-centered interventions aimed at preventing and alleviating suffering in this population.

## INTRODUCTION

Parkinson’s disease (PD) is the second most common neurodegenerative disorder. In 2021, it was estimated to affect approximately 12 million individuals worldwide, a number projected to rise to 25 million by 2050.^1^ Despite its growing impact, various distressing symptoms, such as pain, depression, and constipation, remain underassessed and undertreated.^2,3^ Additionally, psychosocial, spiritual, practical, and carer burden-related concerns are often underestimated. Notably, the provision of palliative care to People Living with PD (PLwPD) is often delayed or entirely absent.^4,5^ PD is also associated with an increased risk of developing dementia, a condition that introduces significant challenges to end-of-life care due to the progressive loss of patient autonomy in decision-making.^6,7^ Recently, the need for greater investment in palliative care for this population has gained recognition, with documented benefits including enhanced quality of life, improved symptom management, and strengthened support for carers.^8,9^

The planning of end-of-life care and the development of new models of care for PLwPD reflect a growing commitment to improving the overall quality of care for this population. Currently, several studies have explored the concept of a ’good death’ from the patients’ perspective across various diseases and the results of these research endeavors have positively influenced the delivery of end-of-life care for these groups.^10–13^ Despite increasing recognition of the importance of palliative care in PD, no studies to date have specifically investigated what constitutes a good death from the perspective of PLwPD.

Given this scarcity of data and the importance of such understanding for providing appropriate palliative care, the present study aimed to give voice to these individuals regarding this topic through a qualitative approach.

## 2. METHODS

This study employed a cross-sectional qualitative design grounded in the constructionist paradigm, which posits that reality is socially constructed and shaped by individuals’ beliefs, practices, and interactions within society.^14^ Inductive thematic analysis was chosen as the analytical method due to its theoretical flexibility and suitability for exploring the perspectives of individuals.^15^

### 2.1 Standard protocol approvals, registrations, and patient consents

All participants had their decision-making capacity to provide informed consent assessed concurrently with the presentation of the Informed Consent Form, following the guidelines proposed by Sudore et al^16^ and previously applied in studies involving interviews on individual values and preferences regarding end-of-life care.^17^

The study was approved by the Ethics Research Committees of the Botucatu Medical School (CAAE: 36453420.1.0000.5411) and the Paulista School of Medicine (CAAE: 36453420.1.3001.5505). In preparing this report, we followed the COnsolidated criteria for REporting Qualitative research (COREQ) guidelines (Appendix 1).^18^

### 2.2 Sample

We used purposive sampling aimed at achieving maximum diversity in age, race, educational level/social class, religious affiliation, and stage of PD among participants from four different health services (two specialized movement disorders clinics, a tertiary geriatric clinic, and a primary care geriatric clinic) in two different cities in the state of São Paulo, Brazil: Botucatu and São Paulo. Participants were initially invited in person by healthcare professionals working in those services. When potential participants agreed to have their contact information shared with the research team, they were contacted by phone to receive further information and an invitation to participate. The sample size was determined based on the concept of Information Power, a model used to guide sample adequacy in qualitative studies considering factors such as the study’s aim, sample specificity, theoretical framework, quality of dialogue, and analytical strategy.^19^ Participant recruitment was discontinued once the data collected were sufficiently rich and diverse to yield nuanced insights and adequately address the study’s objectives, without the need for additional data to enhance meaning or interpretation.

### 2.3 The interview procedure

Interviews were conducted between May 2021 and December 2022, using a semi-structured topic guide (Appendix 2) developed by the research team based on a previous study.^20^ The interview guide was pilot tested with one PLwPD before the start of the study. Depending on logistical considerations, such as participant preference and geographic distance, interviews were conducted either in person at participants’ homes (audio-recorded) or remotely via the Google Meet platform (audio- and video-recorded). Field notes were made after the interviews whenever appropriate. Interviews were conducted by three researchers trained in qualitative methods not hitherto known to the participants: a male physical therapist (LM) with extensive experience working with PLwPD, a female (JMV) and a male medical students (RSC), both with limited experience working with PLwPD.

Following the interviews, sociodemographic data were verbally collected, including age, self-reported gender and race, years of education, approximate time since diagnosis, religious affiliation, relationship with the primary carer, and whether the carer cohabited with the participant. Data on PD staging were obtained using the Hoehn & Yahr classification system, based on participant responses regarding mobility, balance, disability, and the presence of unilateral or bilateral parkinsonian symptoms.^21^ Participants were accordingly classified from stage 1 (unilateral involvement with minimal or no functional disability) to stage 5 (confined to bed or wheelchair unless aided).

### 2.4 Data analysis

The content of the interviews was transcribed verbatim and analyzed using inductive thematic analysis, a method that involves identifying, analyzing, and reporting recurring patterns across the data. The process followed six stages: 1) familiarization with the data, 2) generation of initial codes, 3) identification of themes, 4) review of themes, 5) definition and refinement of thematic narratives, and 6) production of the final report.^22^ We used the Taguette software during the coding process of the thematic analysis.^23^ Three researchers (LM, RSC, and EIOV) coded the data independently, then met to discuss, refine and standardize coding approaches.

### 2.5 Reflexivity

Our research team was composed of professionals from diverse clinical and academic backgrounds, including physical therapy, nursing, medicine (geriatrics, palliative care, pain medicine, and neurology), and psychology, as well as two medical students. We recognized that this dominant clinical positionality inclined us toward a "problem-solving" lens, potentially leading us to over-emphasize patient deficits. To address this, we used our multidisciplinary analysis sessions not just for triangulation, but for active reflexivity, where we systematically challenged one another’s clinical assumptions.

## 3 RESULTS

We conducted 30 interviews, 13 of which were held in participants’ homes, while the remaining 17 were conducted online. Three people who initially had shown interest in the study later declined to participate or did not respond to follow-up contacts. The median interview duration was 27 minutes, with a range from 17 to 164 minutes. All but eight interviews had a carer or family member present based on the participants’ preferences or needs. All participants were interviewed a single time.

The sample was diverse, comprising PLwPD of varying ages, educational backgrounds, religious affiliations, time since diagnosis, and disease stages (Table 1). Although death is often regarded as a taboo subject, participants appeared comfortable discussing it during the interviews.

**Table 1.** Characteristics of participants.

| Characteristic | N (%) or Median<br>(Range) |
| --- | --- |
| Age, median (range) | 67 (36 to 84) |
| Gender, N (%) |  |
| Female | 11 (37%) |
| Male | 19 (63%) |
| Race, N (%) |  |
| Black | 5 (17%) |
| White | 25 (83%) |
| Years of education, median (range) | 8 (1 to 20) |
| Time since diagnosis in years, median (range) | 7 (3 to 40) |
| Stage of Parkinson's Disease, N (%) |  |
| Stage 1 | 5 (17%) |
| Stage 2 | 5 (17%) |
| Stage 3 | 10 (33%) |
| Stage 4 | 9 (30%) |
| Stage 5 | 1 (3%) |
| Relationship with primary carer, N (%) |  |
| Spouse | 2 (7%) |
| Daughter | 1 (3%) |
| Son | 4 (13%) |
| Daughter-in-law | 1 (3%) |
| Friend | 1 (3%) |
| Does not need a carer | 21 (70%) |
| Carer lives with participant | 7 (23%) |
| Religious affiliation, N (%) |  |
| Catholicism | 18 (60%) |
| Evangelicalism | 9 (30%) |
| Spiritism | 2 (7%) |
| Buddhism | 1 (3%) |

We identified two major themes from the data, reflecting complementary dimensions of the concept: *Fears* and *Coping*. Each theme encompassed several subthemes, which captured specific fears and dimensions of coping (Figure 1).

**Figure 1:**
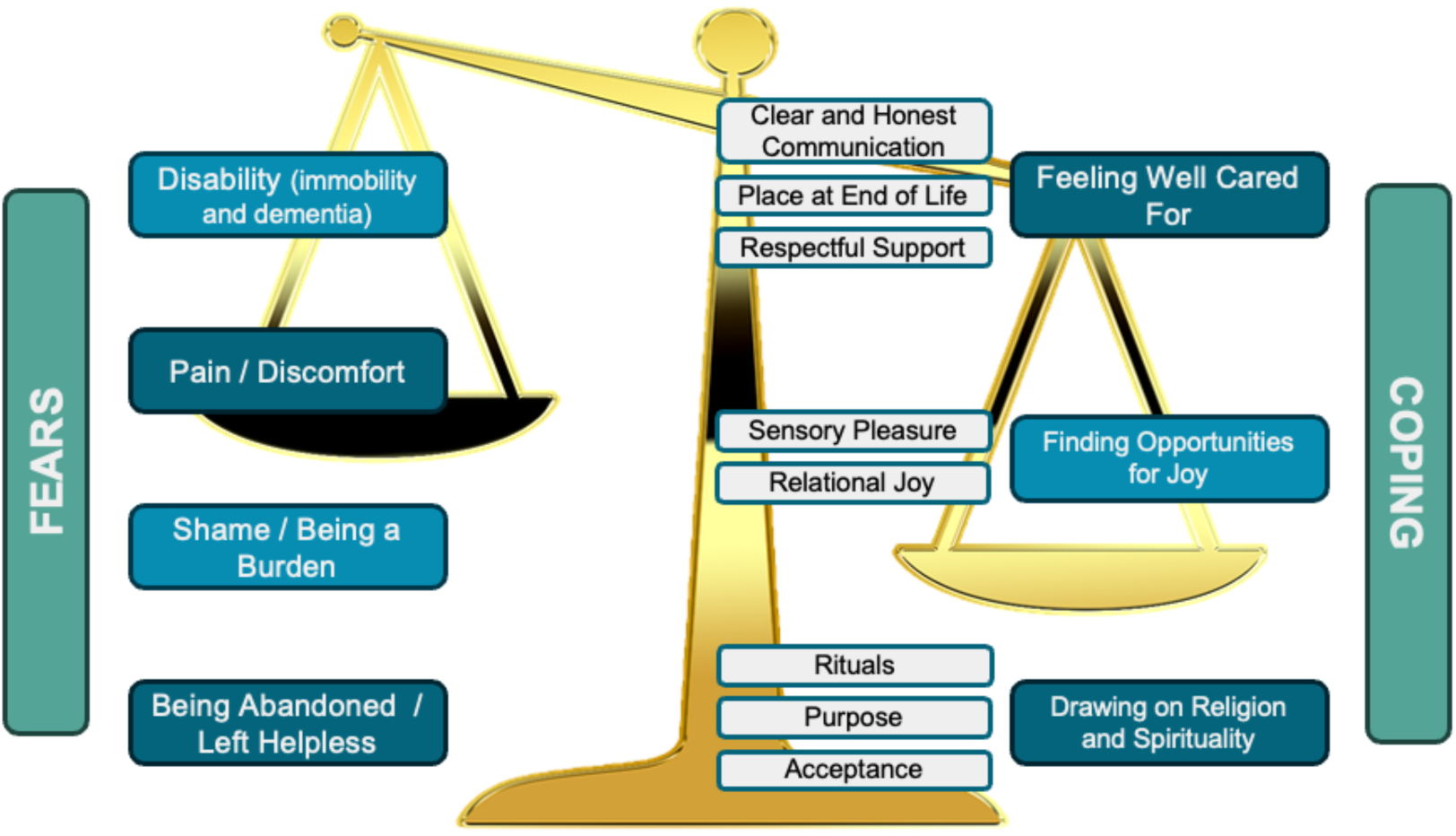
Thematic model derived from qualitative interviews with people living with Parkinson’s disease, showing two main themes: Fears and Coping. The image uses the metaphor of a balance scale to represent how achieving a good death involves counterbalancing fears with coping mechanisms.

### 3.1 Fears

Fear played a central role in how participants articulated their views on the end of life. It manifested across a range of dimensions, encompassing both physical and psychosocial concerns. These included experiences related to their own disease trajectory, observations of others living with PD or facing end-of-life situations, as well as imagined scenarios. Based on thematic analysis, these fears were grouped into four subthemes: (1) fear of disability; (2) fear of pain and discomfort; (3) fear of shame and of being a burden; and (4) fear of abandonment and helplessness.

#### 3.1.1. The fear of disability

Participants often expressed fears related to the anticipated progression of their disease and the consequent loss of independence. Imagining a future marked by continuous struggle often provoked anxiety. The prospect of losing occupational, social, motor, mental, and self-care abilities was perceived as a profound threat to one’s identity and dignity as an autonomous individual.

> *“From what I’ve seen… I think… I’m going to… be, be bedridden… needing… to stay… with almost no movement at all, because I’m losing my mobility, I’m noticing… and needing someone to… turn me to the side in bed, to sit me up, to feed me… I’m, I’m already foreseeing all of this!… I’m (…) trying to come to terms with… what lies ahead, because I know I’m going to die from this… […] I got kinda… yeah, thoughtful when the doctor asked my son, ’Does he still [emphasized the word ’still’] do his business on his own? Like, use the toilet, on his own?’ So… I’m thinking that this will happen to me later on, the way he asked this question to my son… I felt a bit upset, you know? Because… he didn’t even tell me… because… he asked… my son, in his way… ’still,’ this ’still’ [emphasized the word ’still’]… I thought, ’Wow, I’ll have to go through all of this!’ “ (P11)*

Confronted with fears regarding disease progression and the resulting loss of functionality and independence, one participant expressed a desire for euthanasia, while another contemplated suicide as a means of preempting future suffering. Interestingly, when euthanasia was mentioned, it was framed within a paternalistic context, wherein the responsibility for such a decision was attributed to the physician rather than the patient.

> *“…about the way I die… you don’t need… I don’t need to wait until I’m completely debilitated, right? I can die before I get to that point […] It’s just that we… we just watch, you know? Like spectators watching the story… and I find that so awful, just… in such bad taste! Yeah… watching the decline, watching it go on, and on, and on… and then suddenly, it ends in a really bad way, with so much suffering… […] Ugh… suffering in relation to everything… the limitations we have to face… your body not obeying you, your body just… suffering… all the stuff that comes with… with the illness […] I find it unfortunate to live in a country that doesn’t have euthanasia. Oh, euthanasia is so necessary, isn’t it? I’ve talked to… to some people during my life, in Europe, with people who said, “You know, in your country I don’t know, but in mine… there are many doctors who decide to end a life, be it a child or an adult, whatever… when they realize there’s no way out, they end it.” (P14)*

#### 3.1.2. The fear of shame and being a burden

Another facet of the fears expressed by participants involved how they perceived their disease progression through the eyes of others. For example, some participants expressed fears regarding the prospect of becoming unable to perform their personal hygiene autonomously and eventually needing a carer or family member to assist them. This resulted in a complex mix of fear, embarrassment, and shame. Such extreme dependence was perceived as the ultimate stage of autonomy loss, where modesty and dignity could no longer be preserved.

> *“I think like this, being in bed depending on others, just for someone to clean me up, that’s the only thing I don’t agree with… I could die right away without going through this situation… I just don’t want my husband to come and clean my butt… That’s my concern, it’s embarrassing… it just… it bothers me only in that aspect… depending on others, for me… as for the rest, I’ll manage, take it in stride.” (P04)*

Beyond the dependence on care, concern for the well-being of loved ones often involved fears related to their emotional suffering. Frequently, the wish for a swift death or to be alone at the time of death was primarily motivated by a desire to spare family members from both emotional distress and the burden of caregiving.

> *“No, I’m not afraid… not even of death, I’m not afraid! [My fear] is this, being in a bed, suffering there, causing worry for those in the house, and… and I wouldn’t want to have… I would want, if it were my request, a quick death!” (P13)*

> *“[To die] without anyone seeing, right? To die and, when you look, you’re already resting there! … The best thing! Because then you don’t suffer much, right? Can you imagine? You’re dying and the person beside you? Can you imagine them seeing you in your final moments there? It’s agonizing, isn’t it?” (P29)*

#### 3.1.3. The fear of pain and discomfort

Participants also feared the occurrence of several symptoms at the end of life. Some of those fears were based on previous experiences related to PD, such as pain, whereas some fears were unrelated to PD and stemmed from prior experiences with the illness of others, such as shortness of breath. Participants also expressed fears regarding the occurrence of various end-of-life symptoms. Some of these concerns were rooted in personal experiences with PD, such as pain, while others stemmed from witnessing the illness of others and were unrelated to PD, for example, the fear of experiencing shortness of breath.

> *“I guess pain is the toughest part, isn’t it? If we have pain… I think the most important thing is pain, right? What I’m afraid of, doctor, is suffering! [laughs] I have a lot of pain sometimes… Now, if a person becomes bedridden, then… Being bedridden, I think, I don’t know, it’s very tough, isn’t it? It’s not easy, no!” (P06)*

> *“It must be really awful to die from not being able to breathe! My dad has asthma, you know, and I’ve seen him have really bad attacks where he couldn’t breathe… It’s really terrible, right? The person is begging for help, you know, asking for help with their eyes, right?” (P08)*

#### 3.1.4. The fear of being abandoned and left helpless

In the anticipated context of progressive loss of autonomy and increasing burden to carers, some participants expressed fears of being abandoned by their family members. This imagined abandonment was associated with a state of extreme vulnerability and accompanied by a sense of uselessness. The suffering linked to this prospect of abandonment manifested on two levels: one rooted in a feeling of injustice, and the other in a perceived loss of self-worth.

> *“I do have a fear, a fear of progressing and ending up without, without… without anyone to take care of me… like if I end up in a wheelchair all the time, becoming helpless in bed like that…” (P03)*

> *“I just want my relatives to understand this… and… from the moment it starts to, you know, tremble, if I ever get there… right?… that people understand that this is not, it’s not a, it’s not a procedure of the, of the, of the… disease, (…) so… The hard part is… people who end up like this, and the family simply abandons them… right? So, a person who worked their whole life… right… when they become someone useless, let’s put it that way, not a useless person, but… not fit for anything, not even for their… for their own thoughts… People abandon them and just let it go… that’s terrible, isn’t it?” (P07)*

### 3.2 Coping

Almost as a mirror image of participants’ fears, we derived a multidimensional coping theme, comprising the relational experience of feeling well cared for alongside the active strategies of finding opportunities for joy and drawing on religiosity and spirituality.

#### 3.2.1. Feeling well cared for

> *‘Feeling well cared for’ synthesized various elements through which participants perceived their fears could be alleviated. It involved three subthemes related to the place of death, the characteristics of the care provided by health and care professionals, and the relationship with their families.*Most participants expressed a preference for dying at home, as this setting embodied the symbolism of home, i.e., a space associated of safety, values, and affection. However, dying in a hospital environment emerged as an acceptable alternative in the face of the possibility of better symptom control, thereby minimizing suffering through appropriate therapies and ensuring comfort and dignity in a critical context.

> *“Ah, [I’d rather die] in my sleep, right?! And not wake up anymore… In our own home, right? Yeah, you sleep and wake up dead. Because it’s the best place in our home, right? There may be nothing there, but there’s no place like home!” (P04)*

> ***“****Yeah, I’d like to die in a hospital!… Oh, suddenly you’re in a hospital, you’re being taken care of, but you’re not going to make it, you know?… or maybe, so I don’t have so much suffering.” (P28)*

Health and care professionals played an important role in comprehensive care. Participants expressed a desire for support, attention, respect, honest communication, and, interestingly, affection, combined with continuity of care, as factors that would foster a sense of security.

> *“Oh, [healthcare professionals can help] by embracing the patient in both their ethical and professional roles, with a lot of love?” (P06)*

> *“They [the healthcare professionals] must tell the truth to the person… this is what they have to do! […] Sometimes they know that the person will not make it, then it will prepare the person, right?” (P25)*

A quality relationship with families was considered a central component of a good death, as participants believe that fear and suffering could be mitigated through love and connection. Importantly, for some participants, there was also a tension between the desire for closeness with family and the wish to protect them from the distress of witnessing their death.

> *“I wish there were someone… The children… I would die more… peacefully, right?… I don’t know if (death will be) peaceful… for me… [their presence] is a demonstration… of love, right?” (P11)*

> ***“****I don’t really want anything in particular [for my last hours of life]. Not even having family around or anything, ’cause… you know, watching someone die isn’t easy. It’s a heavy thing, especially when it’s someone you love. Makes it even worse. So yeah…” (P07)*

#### 3.2.2. Finding opportunities for joy

Some participants expressed a desire for opportunities to experience joy in the face of their final moments of life. These wishes took various forms, ranging from sensory and gustatory experiences to engaging in specific activities such as traveling, pursuing hobbies, and even shopping. A common feature across these desires was the shift in focus from dying to living, achieved through identifying potential sources of joy.

> *“I think, oh, don’t let me run out of my cigarettes, and don’t let me run out of my sweets! Chocolate!! Chocolate, especially! [It would be] ideal!” (P08)*

> *“I really love to travel, you have no idea! To keep traveling. If I could die, if I could die while traveling. Yeah, the way I like it! It’s a wish, because I love to travel!” (P04)*

In addition to the wishes for sensory sources of joy mentioned above, participants also expressed a desire for relational joy, which arises through interactions with loved ones. Beyond the previously noted reassuring and alleviating role of family during the last moments of life, interviewees emphasized the presence of family members as a source of happiness at the end of life, perhaps reflecting a longing to relive, even briefly, the emotional warmth and joy experienced in earlier shared moments, such as family gatherings during celebrations.

> *“That the whole family would come together, but I don’t know if it’s possible. Before I die, right? I wanted to see everything like that, well, right? […], my wish was that, you know? I always think about that… Then I would leave very happy.” (P17)*

#### 3.2.3. Drawing on religiosity and spirituality

The theme of religiosity and spiritualty encompassed desires for practices that foster some form of spiritual dialogue, such as the anointing of the sick and confession with religious representatives. These practices were regarded as highly meaningful, serving as mechanisms for emotional regulation by addressing fears related to suffering and uncertainty. They represented pathways to spiritual comfort, strength to face adversity and the transition to death, as well as a sense of clear conscience, for example, through the forgiveness of sins.

> *“No. No, I just thought, on the day of my death, that my wife or anyone else (…) with my nephew… to play Ave Maria by Gounod.” (P07)*

> *“As a Catholic that I am, I, I, I would like to, I really want to, go through a confession, with a priest, right? Receive the sacrament of communion and be prepared to go to God, right? Confession leads me to have forgiveness for my sins…” (P30)*

Religiosity and spirituality also conveyed to participants a sense of purpose and acceptance. Focusing on achieving goals related to their religious beliefs provided a feeling of agency while offering a pathway to peace. At the same time, religious convictions often helped participants come to terms with aspects related to their illness and future death that lay beyond their control. Acknowledging that the circumstances of their deaths were inexorable and determined by God’s will, which, according to their religious views, is inherently good, promoted acceptance and alleviated fears. Importantly, prayer also appeared to promote emotional regulation, while reinforcing a sense of agency and acceptance in the face of death.

> *“Yeah, I’m no better than anyone else, serving God the way I’m serving, right? I’m like this, not totally prepared, but yeah… Happy with the situation I live with Parkinson’s! Because I serve God, and I can’t fear this part […] that’s what’s important, and to have a calm, peaceful death, without fear […] If He wants to take me away! [laughs] I’m already going happy! […] For me, for me it’s a good way! For me it is!”(P02)*

> *“Well, in the end, nobody can do anything!… Nobody can do anything! Just have…’it’s… as they say,’it’s… pray to God that everything goes well… look out for those who stay… and t’at’s it!” (P13)*

Religiosity and spirituality also served as interpretive lenses through which participants made sense of their experiences, enhancing the comfort derived from them. For example, health and care professionals were often perceived as bearers of a divine gift, manifested through their actions and conversations aimed at bringing comfort to those under their care.

> *“[Healthcare professionals] seem to have that gift that God gave them! (…) So what they talk with people… Not all of them, but they are pleasant with the person at these times… at least the ones I know, have always been like that.” (P17)*

## 4 DISCUSSION

This study is the first to explore the concept of a good death from the perspective of PLwPD. Our findings align with prior literature on the concept of a good death in other clinical contexts, while also offering novel insights that are particularly relevant for the provision of palliative care for PLwPD.

Our findings on the concept of a good death for individuals with PD can be illustrated by the image of a scale: on one side, the many fears they experience; on the other, the strategies they employ to cope with and alleviate those fears, as depicted in Figure 1. From this perspective, achieving a good death involves tipping the balance by strengthening coping mechanisms and reducing fears. A key contribution of this study lies in deepening our understanding and sensitivity to these fears and dimensions of coping. Healthcare professionals caring for PLwPD often focus primarily on issues related to the motor and non-motor symptoms of that illness while other dimensions remain hidden in plain sight, a phenomenon akin to *inattentional blindness*, exemplified by the classic 1999 ‘invisible gorilla’ study by Simons and Chabris,^24^ whose striking findings were recently replicated in the medical field among radiologists.^25^ Our results have the potential to enhance clinicians’ awareness of patients’ fears and the coping strategies that may help mitigate them. For instance, some of the fears reported by our participants, such as embarrassment and shame at the prospect of a spouse performing intimate hygiene, fear of being abandoned as someone “useless” or “without value”, and fear of being a source of suffering for loved ones, are rarely identified or addressed in clinical practice.

Several previous studies have explored the concept of a good death from the perspectives of various patient populations. A systematic review examining this concept from the viewpoint of patients included 29 studies involving individuals with serious illnesses, such as cancer, cardiovascular diseases, chronic obstructive pulmonary disease, and HIV/AIDS, as well as older adults and members of the general population.^26^ The review synthesized its findings into six core elements of a good death and six overarching themes that shape this concept. The identified core elements were: effective pain and symptom management; being recognized and treated as a person; clear communication and shared decision-making; the ability to contribute to others; a sense of closure; and preparation for death. The main themes influencing these elements included religion and spirituality, cultural and societal factors, financial concerns, life circumstances, age, and disease. Another recent review focusing on the perspectives of individuals with cancer identified seven key themes: awareness of the disease, management of pain and symptoms, dying well, being remembered after death, personal interpretations of a good death, individual behaviors that contribute to a good death, and the influence of culture and religion.^27^

On the one hand, several elements from our findings align with those reported in previous reviews, as, for example, the fear of pain and discomfort, the wish for honest communication with healthcare professionals, the fear and shame of becoming a burden to loved ones, and the importance of religiosity and spirituality. On the other hand, our study offers additional insights specific to individuals with PD that were addressed in those reviews. In particular, the fear of disability was a central theme for our participants with PD yet was absent from the prior literature. This suggests that concerns about losing mobility and independence are distinctive factors shaping how people with PD think about their end of life, and which require careful attention from healthcare professionals. In contrast to people with cancer, whose diagnosis is often equated with death,^28^ PD is more strongly linked to the notion of disability, which then becomes a central fear when contemplating the end of life.

Another important insight that appeared in our study but was absent from previous reviews concerns the value of creating opportunities for joy. This is a particularly compelling topic, yet it is not addressed in sections devoted to palliative care within influential guidelines on the general management of PD,^29^ nor in guidelines specifically focused on palliative care for PLwPD.^5,9^ The value in identifying and promoting opportunities for joy in the face of serious illnesses has been recognized by palliative care and mental health professionals but often remains poorly integrated into clinical care.^4,30^ Our study serves as a reminder to clinicians caring for people with PD of the significance of promoting joy as part of comprehensive, person-centered care.

Our study has several limitations. First, none of our participants seemed to be in their final days of life. This required them to engage in a certain level of abstraction to imagine their last moments along with the health scenario they envision for the future, within the constraints of uncertainties regarding the progression of PD. Consequently, the perspectives shared by the participants reflect their current fears and anxieties about the future, which may evolve in response to future aggravating factors, such as unexpected disease symptoms, or changes in family dynamics during the course of increasing dependency. However, it would be logistically challenging and ethically questionable to interview people who were actively dying – a challenge that has not been overcome by previous studies on the concept of a good death in other populations. Notably, although we did not track clinical outcomes, we were informed by the families of two participants that they had passed away within a few months of their interviews. Second, the study was conducted exclusively with patients residing in two cities within a single state in one Latin American country. Nevertheless, we employed a multicenter design and recruited participants from four distinct healthcare services. As is typical in qualitative research, our goal was not to produce statistical generalizations but to deepen understanding of what constitutes a good death for individuals living with PD. In qualitative studies, transferability is understood as a collaborative process between researchers and readers.^31,32^ Based on the researchers’ descriptions and interpretations, readers assess the extent to which the findings resonate with and are applicable to other contexts that share similar characteristics. Third, we did not return transcripts to participants for comment or conduct member checking.^33^ Although this practice is increasingly adopted in qualitative interview-based studies, its contribution to credibility remains uncertain, and the process may pose emotional burdens for participants.^34,35^ Lastly, our research team did not include individuals with lived experience of PD, who could have contributed to the development of interview questions and the interpretation of participants’ narratives. Instead, the research protocol and analysis were conducted exclusively by healthcare professionals.

On the other hand, our study presents several strengths. Few studies on this topic have been conducted in middle- and low-income countries, particularly in Latin America.^20^ Our sample included patients at various stages of PD, from four distinct healthcare services, with diverse educational backgrounds, disease durations, and religious affiliations. Importantly, this research benefited from the collaboration of professionals from multiple disciplines, whose varied perspectives enriched the interpretation of participants’ narratives.

Our findings have implications for both clinical practice and research beyond issues related to the end-of-life care of this population. This study invites clinicians to question how they approach support and whose priorities are centered during consultations. While clinicians often focus on motor and non-motor symptoms of PD, our results illustrate that patients are navigating a complex landscape of fears, coping mechanisms, and the pursuit of feasible joy – issues often overlooked in clinical practice. The core implication of our results is the need to shift our clinical lens from the disease to the person faced with progressive decline as they near the end of their life. This requires more than simply adding ‘coping’ to a checklist; it demands centering the patients’ perspectives and priorities in all communication and care planning. It is only from this patient-centered stance that effective, non-pharmacological interventions aimed at alleviating fears and strengthening coping strategies can be meaningfully developed and tested in partnership with the patients.

## 5 CONCLUSION

Our findings suggest that improving palliative care for PLwPD requires an approach that actively addresses specific fears and strengthens the multiple dimensions of coping, which includes fostering opportunities for joy, supporting spirituality, and enhancing the relational experience of feeling well cared for. This study illuminates often-overlooked aspects of care and provides a basis for the development of person-centered interventions aimed at preventing and alleviating suffering in this population.

## Acknowlegments

The authors are very grateful to the participants, who were generous with their time.

## Authors’ contributions

LM and EIOV had full access to all the data in the study and take responsibility for the integrity of the data and the accuracy of the data analysis and are the guarantors for the study. Concept and design: LM and EIOV. Acquisition of data: LM, RSC, and JMV. Analysis: LM, RSC, RM, JMV, DO, HBF, NRT, FBF, and EIOV. Drafting of the manuscript: LM. Critical revision of the manuscript for important intellectual content: RSC, RM, JMV, DO, HBF, NRT, FBF, and EIOV. All authors approved its final version for submission.

## Data availability

Because we do not have the permission of participants to share the transcribed interviews and to preserve their anonymity, the transcribed interviews are not available for sharing. In case other researchers wish to have access to the anonymized interview transcripts they should contact the corresponding author while understanding that any access to the transcripts is conditional to the approval by the specific Ethics Review Committee.

## Appendix 1 COREQ (Consolidated criteria for REporting Qualitative research) Checklist

A checklist of items that should be induded in reports of qualitativerest?arch. Youmust report thepage number in your manuscript where youconsider each of the?items listed in thischecklist. If youhave? not included thisinformation, t?itht?r rnvisC? your manuscript accordingly beforn submitting or note N/A.

| Topic | Item No. | Guide Questions/Description | Reported on<br>Page No. |
| --- | --- | --- | --- |
| **Domain 1: Research team and reflexivity** |  |  |  |
| *Personal characteristics* |  |  |  |
| Interviewer/facilitator | 1 | Which author/s conducted the interview or focus group? | 5 |
| Credentials | 2 | What were the researcher's credentials? E.g. PhD, MD | 5 |
| Occupation | 3 | What was their occupation at the time of the study? | 5 |
| Gender | 4 | Was the researcher male or female? | 5 |
| Experience and training | 5 | What experience or training did the researcher have? | 5 |
| *Relationship with participants* |  |  |  |
| Relationship established | 6 | Was a relationship established prior to study commencement? | 5 |
| Participant knowledge of the interviewer | 7 | What did the participants know about the researcher? e.g. personal goals, reasons for doing the research | 5 |
| Interviewer characteristics | 8 | What characteristics were reported about the interviewer/facilitator? e.g. Bias, assumptions, reasons and interests in the research topic | 5 |
| **Domain 2: Study design** |  |  |  |
| *Theoretical framework* |  |  |  |
| Methodological orientation and Theory | 9 | What methodological orientation was stated to underpin the study? e.g. grounded theory, discourse analysis, ethnography, phenomenology, content analysis | 4 |
| *Participant selection* |  |  |  |
| Sampling | 10 | How were participants selected? e.g. purposive, convenience, consecutive, snowball | 5 |
| Method of approach | 11 | How were participants approached? e.g. face-to-face, telephone, mail, email | 5 |
| Sample size | 12 | How many participants were in the study? | 6 |
| Non-participation | 13 | How many people refused to participate or dropped out? Reasons? | 6 |
| *Setting* |  |  |  |
| Setting of data collection | 14 | Where was the data collected? e.g. home, clinic, workplace | 6 |
| Presence of non-participants | 15 | Was anyone else present besides the participants and researchers? | 6 |
| Description of sample | 16 | What are the important characteristics of the sample? e.g. demographic data, date | Table 1 |
| *Data collection* |  |  |  |
| Interview guide | 17 | Were questions, prompts, guides provided by the authors? Was it pilot tested? | Appendix 2 |
| Repeat interviews | 18 | Were repeat interviews carried out? If yes, how many? | 6 |
| Audio/visual recording | 19 | Did the research use audio or visual recording to collect the data? | 5 |
| Field notes | 20 | Were field notes made during and/or after the interview or focus group? | 5 |
| Duration | 21 | What was the duration of the interviews or focus group? | 6 |
| Data saturation | 22 | Was data saturation discussed? | 5 |
| Transcripts returned | 23 | Were transcripts returned to participants for comment and/or | 16 |

| Topic | Item No. | Guide Questions/Description | Reported on Page No. |
| --- | --- | --- | --- |
|  |  | correction? |  |
| **Domain 3: analysis and findings** |  |  |  |
| *Data analysis* |  |  |  |
| Number of data coders | 24 | How many data coders coded the data? | 6 |
| Description of the coding tree | 25 | Did authors provide a description of the coding tree? | NA |
| Derivation of themes | 26 | Were themes identified in advance or derived from the data? | 4 |
| Software | 27 | What software, if applicable, was used to manage the data? | 6 |
| Participant checking | 28 | Did participants provide feedback on the findings? | 16 |
| *Reporting* |  |  |  |
| Quotations presented | 29 | Were participant quotations presented to illustrate the themes/findings?<br>Was each quotation identified? e.g. participant number | 7-14 |
| Data and findings consistent | 30 | Was there consistency between the data presented and the findings? | 7-14 |
| Clarity of major themes | 31 | Were major themes clearly presented in the findings? | 7-14 |
| Clarity of minor themes | 32 | Is there a description of diverse cases or discussion of minor themes? | 7-17 |

Developed from: Tong A, Sainsbury P, Craig J. Consolidated criteria for reporting qualitative research (COREQ): a 32-item checklist for interviews and focusgroups. *lntemotlonal Joumalfor Quality In Health Care.* 2007.Volume 19, Number 6: pp. 349 – 3S7

Once you have completed this checklist, please savea copy and upload It as part of your submission. DONOTlndude this checklist as part of the mainmanuscript document. It must be uploaded asa separate file.

## Appendix 2 Guide for Semi-Structured Interview

Thank you for agreeing to take part in this interview. In this research, we would like to hear your opinion about what would constitute a good death.

This interview aims to improve our understanding of the needs and preferences of people with Parkinson’s disease, with the goal of enhancing the quality of their end-of-life experiences.

If at any time during the interview you need to stop or leave the room, please let me know.

Everything that you tell me will be treated anonymously, avoiding your exposure.

At the end of the interview, if you have any additional comments or questions that you would like to share, please let me know, so I can record them.

The main goal of this interview is to understand:

- What a good death means to you
- What a bad death means to you
- What kinds of things could make a death “good”

### QUESTIONS

1. First of all, what does a good death mean to you? *Potential prompts*
  - Do you remember having experienced the death of someone close to you in the past?

∘ What was it like? Did the person die from a diagnosed illness? Do you know what it was?
2. Are there specific things that you would wish for as you near death?
3. Are there specific things that you fear for as you near death?
4. Does your Parkinson’s diagnosis make any difference in what a good death means to you?
5. What do you understand about what might happen to you, as Parkinson’s disease progresses?
6. Do you feel you know as much as you want to know about what may happen to you as you near death? *Potential prompts*
  - How important is it to you to know in advance what is likely to happen as you near death?
  - What are your thoughts and feelings about the uncertainties around death and dying?
  - What symptoms do you think people may experience when dying?

∘ *Wait for spontaneous response. If the interviewee asks for examples, mention* pain, difficulty breathing, incontinence, inability to chew or swallow, or loss of consciousness.
7. Is it important for you to have control over your death? [If yes] What would having control feel like? *Potential prompts*
  - What kinds of preparations do you think can be made for a person’s end of life?
  - Have you made any preparations or thought about doing so? (For example, a will, an advance directive, etc.)
8. Would you like to be alone or have someone with you during your final days of life? *Potential prompts*
  - How important would it be to have family or friends nearby? How might the presence of family and friends influence the quality of your death?
  - Would you like to have anyone else nearby, such as a healthcare professional, a representative of your religion (e.g., priest), or a pet?
  - What kind of role would you like them to have?
  - Is it important for you to be able to say goodbye to people? If so, when and how?
9. Where do you think would be a good place to die? *Potential prompts*
  - Do you have a preference for dying in your own home, someone else’s home, or in a hospital? Why?
  - Is there any place where you definitely would not want to die? Why?
  - If dying in a hospital meant that your symptoms (for example, pain) would be better controlled, would that affect your choice of where to die and why?
10. How can health and social care professionals influence the quality of your death? *Potential prompts*
  - And what about nurses, doctors, and formal caregivers? Anyone else?

